# A medically grounded LLM agent–based tool to detect patient safety events in medical records

**DOI:** 10.64898/2025.12.16.25342438

**Authors:** Diego Trujillo, Dulin Wang, Nathan Bahr, Tina Yi-Jin Hsieh, Byeongyeon Cho, Garth Meckler, Matthew Hansen, Carl Eriksson, Kyu Seo Kim, Steven Bedrick, Xiaoqian Jiang, Jeanne-Marie Guise

**Affiliations:** Center for Learning Health Care Delivery, Beth Israel Deaconess Medical Center, Boston, Massachusetts; Department of Obstetrics Gynecology and Reproductive Medicine, Beth Israel Deaconess Medical Center and Harvard Medical School, Boston, Massachusetts; McWilliams School of Biomedical Informatics, UTHealth, Houston, Texas; Department of Emergency Medicine, Oregon Health and Science University, Portland, Oregon; Department of Biomedical Informatics, Harvard Medical School, Boston, Massachusetts; Department of Pediatrics and Emergency Medicine, University of British Columbia, Vancouver, Canada; Department of Pediatrics, Oregon Health and Science University, Portland, Oregon; Department of Medical Informatics and Clinical Epidemiology, Oregon Health and Science University, Portland, Oregon

## Abstract

Large language models (LLMs) have shown incredible promise in medicine. While LLMs may be particularly useful in areas requiring extensive review of clinical records, their use remains limited due to their tendency to hallucinate and fabricate information. Hallucination issues, as well as their consequences, are exacerbated in low–probability, high–stakes scenarios such as rare adverse safety events or medical errors. We present SAFE–AI (Structured and Automated Framework for Explainable AI), a novel method for clinical decision making that combines the strengths of clinical expert knowledge with LLMs in an ontology–driven model that minimizes hallucinations using strict rules. We test this method to identify medication errors in medical charts. We collected a sample of 18,402 lines of clinical information from 300 EMS clinical charts that were independently dually reviewed by two expert physicians for epinephrine adverse safety events (ASEs), with 96% inter-rater agreement. We tested SAFE–AI against these labels, achieving human–like performance in detecting epinephrine overdoses with 97.9% accuracy, and 91.6% accuracy in identifying delays in epinephrine administration, greatly outperforming baseline LLMs models. Notably, some disagreements between clinicians and the model were found to be justifiable differences in judgment rather than errors. SAFE-AI presents a novel approach for clinical AI applications that addresses two key limitations of current machine learning methods: 1) over-reliance on probabilistic pattern recognition instead of established medical knowledge, and 2) perpetuation of biases present in training data. This framework is easily adaptable to a range of clinical applications, paving the way for provable and trustworthy AI in medicine.

**Author Summary:** LLMs have shown promise in analyzing clinical records but their use is limited due to their tendency to hallucinate and fabricate information. Misinformation could threaten patient safety and jeopardize trust. We developed SAFE–AI (Structured and Automated Framework for Explainable AI), which combines knowledge from clinical experts with LLM inference to detect adverse safety events (ASEs) with minimal errors. We tested our method in identifying medication errors in medical charts and compared results to reviews by expert physicians. Our method detected epinephrine delays and overdoses with a high level of accuracy. SAFE-AI presents a novel approach for clinical AI applications that overcomes reliance on pattern recognition instead of medical knowledge biases present in training data.

## Introduction

It is estimated that over 250,000 adverse safety events (ASEs) occur annually in the United States among patients receiving medical care, resulting in more than 100,000 patient deaths [1, 2, 3]. ASEs, such as medication, diagnostic, or procedural errors; miscommunications; and healthcare-associated infections, pose significant risks to patient safety in healthcare settings. The current standard for detecting ASEs is for expert clinicians to manually review clinical records [4, 5]. Manual chart reviews are labor-intensive, slow, and costly. Consequently, ASEs are undetected and under-reported, with over 95% of documented ASEs in medical records not being included in the official records, which hampers quality improvement and patient safety efforts [6].

Recently, artificial intelligence (AI) and particularly large language models (LLMs), have shown promise for aug-menting healthcare professionals’ abilities to analyze complex medical data. However, these models have limitations such as the propensity for LLMs to “hallucinate” – giving fabricated or fictional information that can be misleading and dangerous in clinical settings [7, 8, 9]. Recent studies have shown that their performance is highly conditional on the probability of the correct outcome [10]. As ASEs are relatively uncommon events, LLMs alone are not likely to correctly identify them. Even more troubling are some recent analyses of LLM robustness, showing the results of LLM requests are greatly dependent on the specific phrasing used [11].

LLM hallucination is most evident when the model is asked to perform reasoning-based tasks, but is greatly mitigated when limited to extracting information. This is demonstrated by the success of techniques such as retrieval-augmented generation (RAG) [12], where the prompt is augmented with additional context from relevant databases before querying the LLM. retrieval-augmented generation (RAG) improves LLM performance and reduces hallucinations by up to 26% [13]. By including context, and potentially the solution, in the prompt, the model acts as an information retrieval system. However, while RAG is successful in improving the results of LLMs, it does not offer any true protection against hallucinations, and still behaves as a black box, lacking explainability and verifiability.

To address these challenges, we developed SAFE–AI (Structured and Automated Framework for Explainable AI), an automated decision-making framework that merges state-of-the-art AI models with structured clinical knowledge. By combining the capabilities of LLMss with dynamic prompting and automatic decision–making based on deterministic logic, SAFE–AI offers fast and reliable identification of ASEs from both free-text clinical notes and structured data sources. Our approach seeks to bridge the gap between clinician knowledge and fast, automatic ASE detection. Our objective was to test the ability of SAFE-AI in reviewing EMS charts for medication errors, specifically potentially lethal overdoses or delays in administering Epinephrine in pediatric out-of-hospital cardiac arrest (OHCA) care [14]. We test this approach on epinephrine dosing and timing – two serious ASEs in – and compare against baseline LLMs and human reviewers.

## Methods

### Overview

SAFE–AI is structured in four major steps (Figure 1). Firstly, an ontology of epinephrine errors is defined by clinician experts using established clinical guidelines, and this ontology is converted into a directed graph. Secondly, an LLM is used to retrieve initial nodes in the graph – such as patient age and weight – from an unstructured patient chart. Thirdly, a coding LLM translates the guidelines defined in the ontology into executable code. Lastly, a final prediction is made by executing the computational graph of all nodes from the initial inputs retrieved by the LLM to the final output node. Thus, the only source of hallucination stems from the initial information extraction - a far simpler task for LLMs to perform accurately.

**Figure 1:**
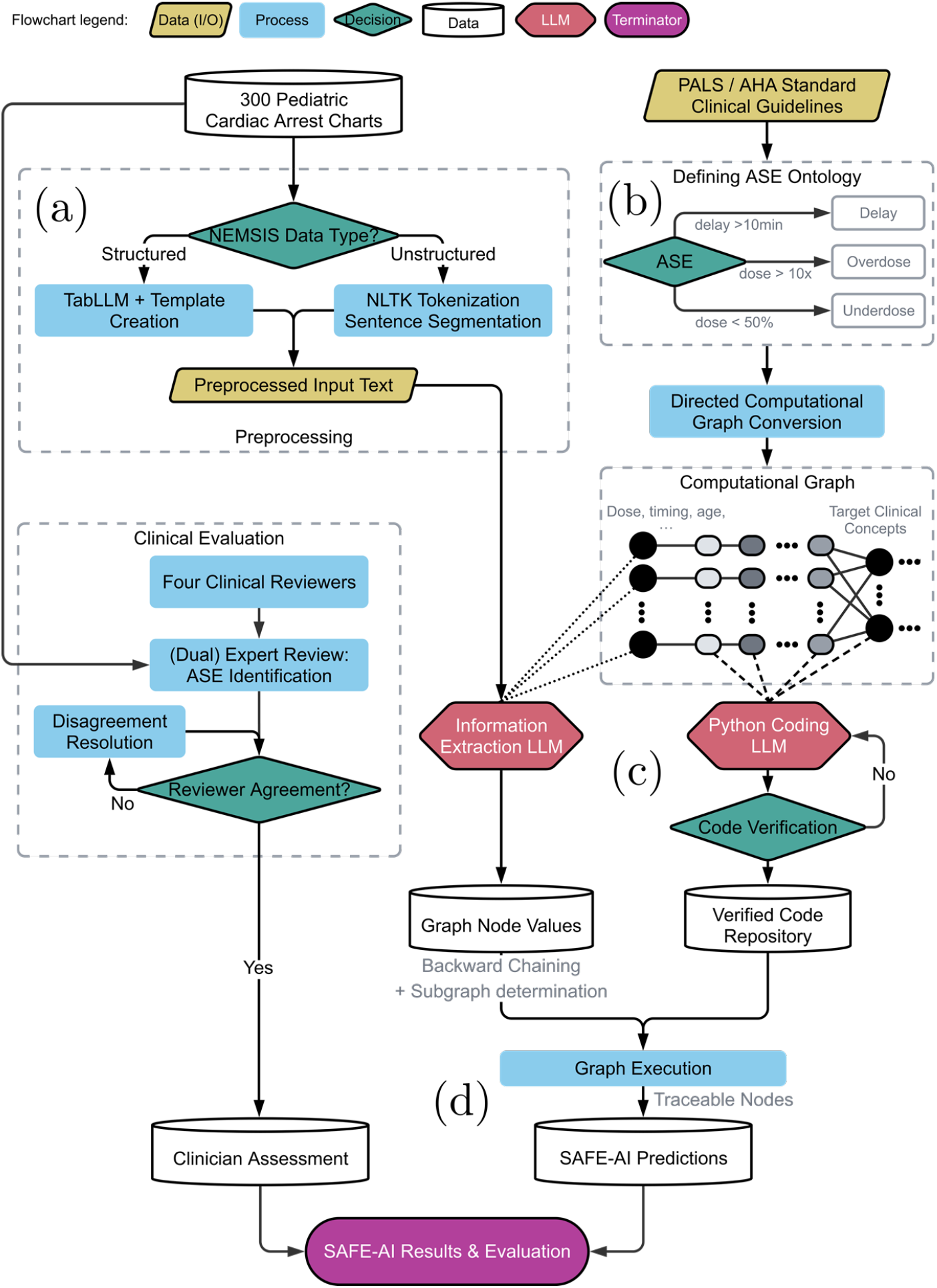
SAFE–AI process diagram. a) An emergency medical services (EMS) chart containing both structured and unstructured information is taken as input; b) an ontology of ASE–related concepts is defined, also defining relationships between concepts; c) inputs to the ontology are extracted from the text and a coding LLM translates the guidelines into executable code for the intermediate nodes in the ontology; d) the complete program makes a final determination by executing the graph with the extracted inputs.^1^

### Data Collection

We acquired a sample of 5089 pediatric OHCA patient care charts from 2014 to 2019 from a large nationwide provider of EMS transports using the National Emergency Medical Services Information System (NEMSIS) standard [15]. The study was approved by Beth Israel Deaconess Medical Center’s Institutional Review Board as exempt. We randomly selected 409 charts to build a gold standard and test our system’s performance. Inclusion/exclusion criteria are shown in Figure 2. The final sample contained 300 charts and 18,402 lines of clinical information.

**Figure 2:**
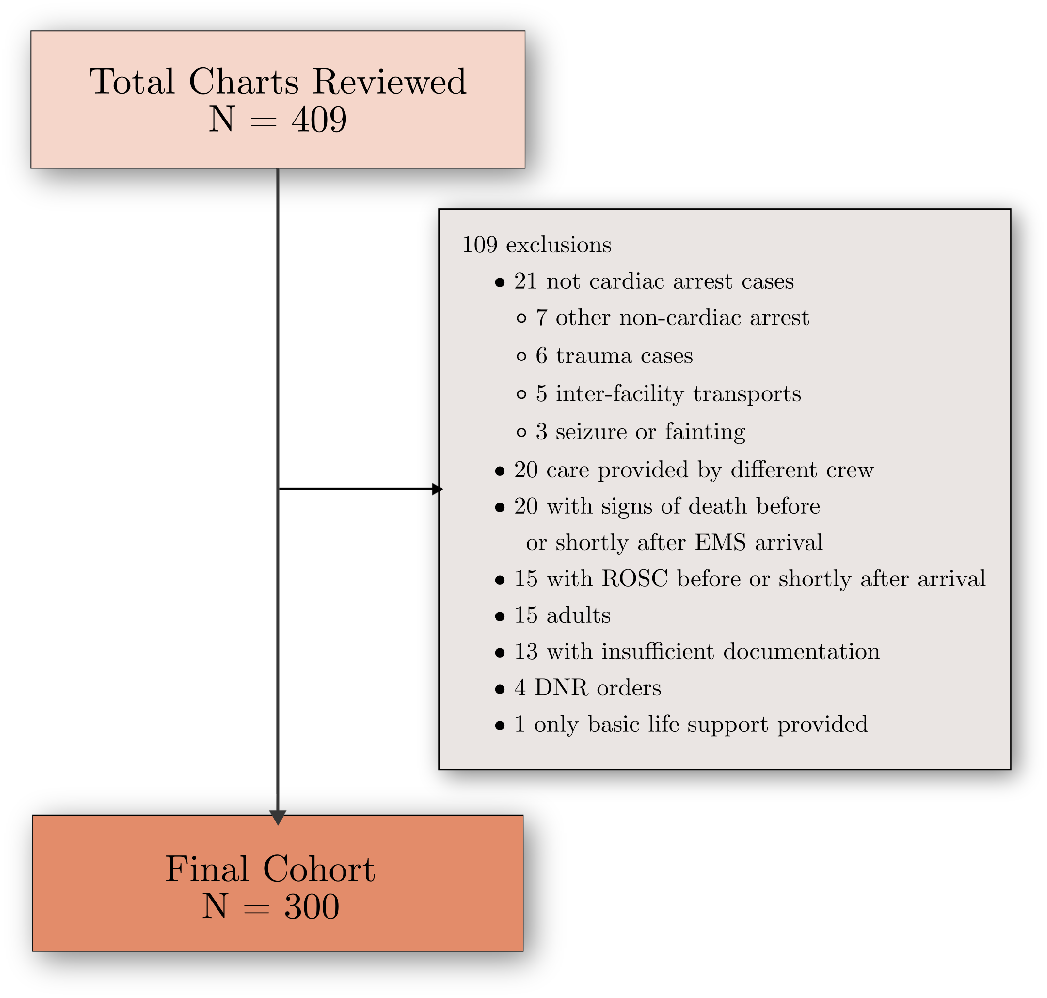
Excluded and reviewed charts.

Clinical experts in pediatric Emergency Medicine and Critical Care independently verified inclusion status and determined presence of ASEs. Following a process described in previous studies [16, 14, 17], board-certified clinical experts used a validated PEDS tool to independently review medical records for ASEs. Reviewers underwent rigorous training and were required to have at least 80% agreement before they were allowed to independently review. Every chart was randomly assigned to two independent reviewers and disagreements were resolved by complete consensus.

We used Gwet’s AC1 to measure inter-rater agreement for epinephrine ASEs [18]. We have used this measure in previous studies [19] to overcome the “kappa paradox” [20], wherein the low prevalence of events, such as severe ASEs, could result in low scores despite high agreement among raters [18, 21, 22].

### Data preprocessing

Charts in the National Emergency Medical Services Information System (NEMSIS) standard contain unstructured narratives and structured fields [15]. We used manually-crafted templates, such as “the patient weighs X kg”, to generate sentences from structured fields, following the strategy used by TabLLM [23]. Unstructured narratives were tokenized into sentences with Python’s Natural Language Toolkit [24]. The tokenized and templated sentences were combined into a single document and used as input for the LLM to perform information extraction.

### Ontology of epinephrine errors

To demonstrate the capabilities of the SAFE–AI framework, we created an ontology to formalize epinephrine ASEs. In previous work [17, 16] we created a classification system that defined epinephrine ASE as failure to administer when indicated, a delay to administer epinephrine within 10 minutes of arrival, and administering at least a 10x overdose or 50% underdose. In this work, we focused on the latter three. To build an ontology, we expanded these definitions with explicit criteria for prerequisites, and constraints.

Epinephrine is indicated in all pediatric pulseless cardiac arrest according to Pediatric Advanced Life Support (PALS) guidelines published by the American Heart Association (AHA) [25]. Using expert consensus and rules developed in prior studies, we developed a specific ontology including specific time thresholds and erroneous doses that represent ASEs with the potential to negatively impact patient outcomes. We defined thresholds for severe ASE, mild ASE, and no ASE (Table 1), and included the generated ontologies for overdose, underdose, and delay in administration (Figure 1).

**Table 1:**
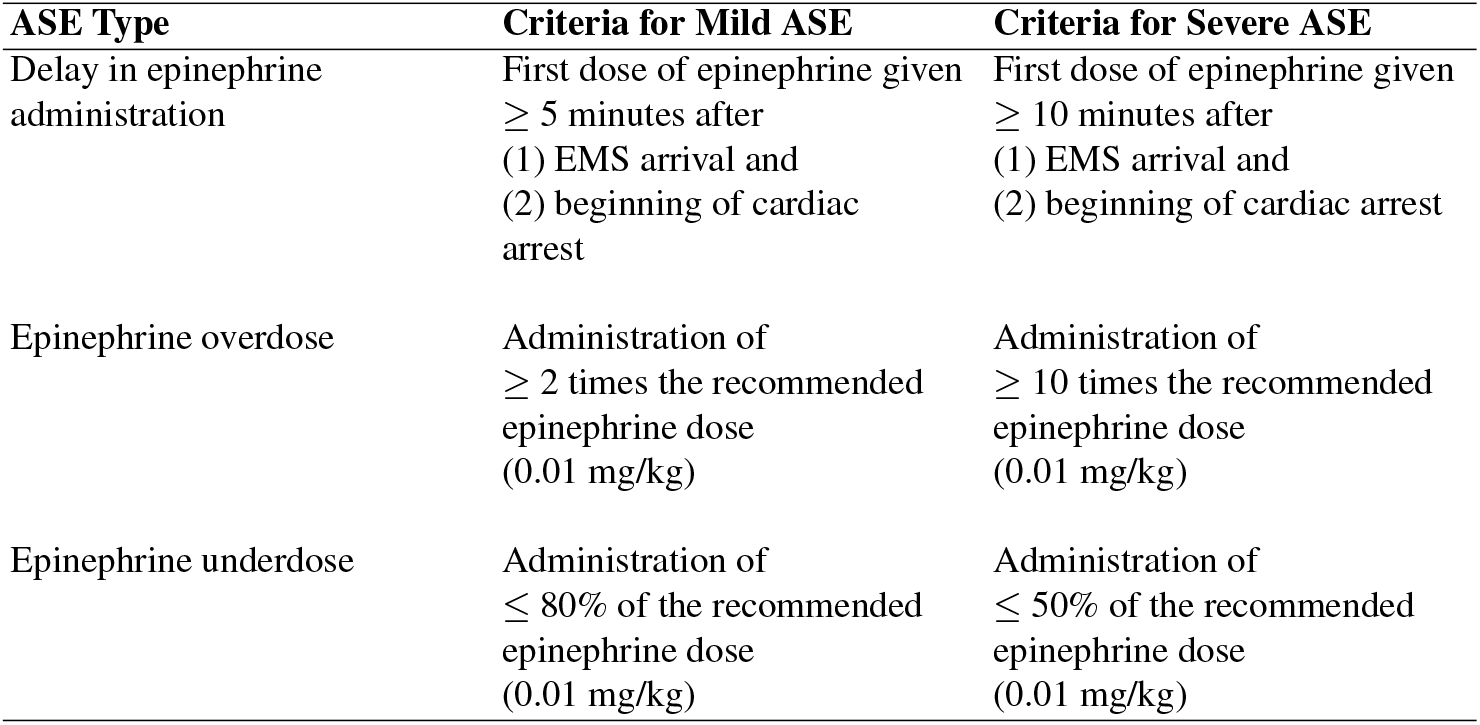
Criteria for severe epinephrine adverse safety events (ASEs).

### LLMs as zero–shot information extractors

The ability of LLMs, such as OpenAI’s GPT–4 [26], to extract information from a text is of special interest in this work, illustrated with the concepts of frequency and recency bias.

Formally, let *C* = (*w*_1_, *w*_2_, …, *w*_*n*_) represent the context, a sequence of tokens; and let *V* be the vocabulary of all possible tokens.

#### Frequency bias

The probability of a token is influenced by its overall frequency in the training data. Specifically, if we define *f* (*w*) as the frequency count of token *w* in the training corpus, then the marginal probability of *w* can be approximated by

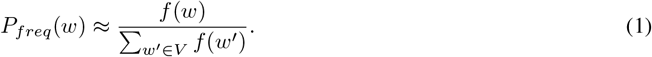

As a result, tokens with higher frequencies have higher baseline probabilities.

#### Recency bias

The attention mechanism in transformer models allows the model to weigh recent tokens more heavily. Due to positional encoding and the design of attention layers, tokens closer to the last position often receive higher attention weights. The most widely used positional encodings (such as the sinusoidal embeddings in the Transformer and RoFormer embeddings [28]) have this property.

These two biases combined show that information extraction is a fundamentally easier task for LLMs than reasoning, as the information often exists verbatim or with slight paraphrasing in the context *C*. In contrast, reasoning often involves generating tokens or sequences that may be low frequency or novel, thus having lower learned probabilities. For this reason, we looked at LLMs as zero–shot information extractors only. We built a pipeline that uses the language model only to retrieve relevant concepts from the ontology of epinephrine errors in Table 1. In doing this, we take advantage of the LLM capabilities to parse unstructured text and retrieve relevant information, such as patient age and weight, with minimal to no hallucinations.

These two biases show that information extraction is a fundamentally easier task for LLMs than reasoning, as the information often exists verbatim or with slight paraphrasing in the context. In contrast, reasoning often involves generating tokens or sequences that may be low frequency or novel, thus having lower learned probabilities. For this reason, we looked at LLMs as zero–shot information extractors. We built a pipeline that uses the LLM to retrieve relevant concepts from the ontology of epinephrine errors. In doing so, we take advantage of the LLM capabilities to parse unstructured text and retrieve relevant information while minimizing hallucinations.

We combine the ontology created in the previous section with a template system to dynamically generate prompts from the developed ontology and structure the outputs into JSON strings.

### Ontologies as computational graphs

Once the relevant inputs have been extracted from the source text with the LLM, a decision can be made with verifiable logic. Using template-based prompting, a coding model (GPT-4o) is asked to translate clinical guidelines into Python code. The executable Python code is run once in a sandboxed environment to be checked for basic compilation errors or malicious code, and the resulting code can be reviewed and verified by experts. SAFE–AI can also use existing functions instead of generating them; in the case of ASE prediction, we generate the code once and save to a file for reuse after verification.

When trying to determine an ASE, SAFE–AI selects the relevant output node in the graph and determines the required subgraph of relevant nodes with backward chaining. Then, after initial nodes in the graph receive their values from the LLM extraction and are coerced to expected data types, the computational graph is executed from the input nodes to the output node. In case of missing inputs or any errors during execution, the program propagates the error and allows tracing it back its origin.

By treating the ontology as a computational graph where nodes represent concepts –such as patient age, administered doses, or dosing ASEs– we ensure traceability of the decision-making. All the intermediate and output nodes have explicit computations that can be verified for accuracy, and only need to be defined once.

## Results

### Chart Review

A final sample of 300 pediatric emergency medical charts was reviewed by clinician experts. Gwet’s AC1 showed excellent inter–rater agreement among clinical reviewers for evaluating epinephrine ASEs, with values of 0.85 for delays of ≥ 10 minutes to administer epinephrine and 0.96 for incorrect epinephrine doses.

There were very few true underdose cases according to chart reviews (*<* 5), preventing us from reliably calculating underdose accuracy, leading us to omit underdose metrics in our reporting. However, SAFE-AI can predict underdoses using the ontology.

We evaluated the extraction performance for selected variables (age, weight, epinephrine dose, EMS arrival time). The results were as follows:

- Empty Field Errors: these occurred when the LLM identified conflicting values and did not output any value. This happened with the age variable (7/300 cases, 2.33%) and the weight variable (1/300 cases, 0.03%).
- Incorrect value errors: these occurred when the chart contained duplicate or conflicting values for epinephrine dosage (5/300 cases, 1.66%).
- In total, 14 of 300 charts (4.66%) showed extraction failures. Of these, only one was clearly unambiguous and had no conflicting information.

### Prediction Results

To evaluate the contribution of ontological structures and decision-making logic in SAFE–AI, we implemented two ablated baselines: (1) LLM only: relying purely on LLM predictions without ontology and decision-making logic, and (2) LLM+ontology: incorporating ontology structured inputs from LLM without executing decision-making logic. These ablations reveal the extent to which structured decision-making improves prediction accuracy beyond raw next–token generation. Results of these ablations, as well as the full SAFE–AI model, are summarized in Table 2.

**Table 2:**
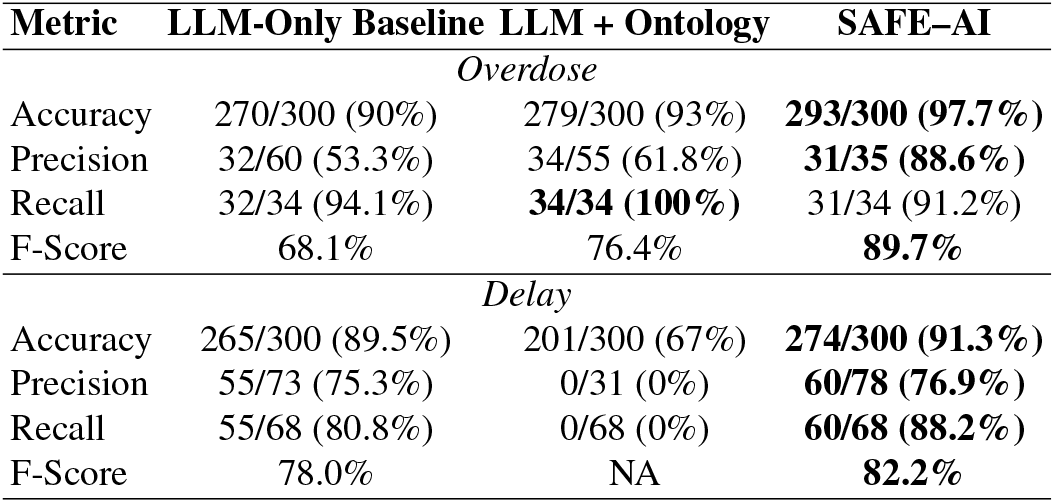
Model results against baselines and ablations.

### LLM-only predictions yield high recall but low precision, leading to inconsistencies

As shown in Table 2, the LLM-Only Baseline achieves a recall of 32/34 (94.1%) for overdose detection and 55/68 (80.8%) for delay detection, correctly identifying most relevant cases. However, its precision remains low (32/60 (53.3%) for overdose, 55/73 (75.3%) for delay), leading to frequent false positives. This results in an F-score of 68.1% for overdose and 78% for delay, demonstrating that while the model captures relevant cases, its lack of structured reasoning leads to inconsistencies in classification.

### Ontological constraints improve overdose detection but severely underperform delay classification

LLM+Ontology improves overdose detection accuracy 279/300 (93%), with precision increasing to 34/55 (61.8%) and recall reaching 34/34 (100%). This results in an improved F-score of 76.4%. However, in delay detection, the model breaks down completely, with an accuracy of 201/300 (67%), and precision and recall of 0. As reported in Table 2, these results suggest that while ontological constraints help maintain logical consistency in structured tasks, they impose excessive rigidity to a prediction.

### SAFE–AI achieves the highest precision and F-score, balancing recall and decision accuracy

SAFE–AI outperforms both ablated models across all metrics, as summarized in Table 2. For overdose detection, it achieves an accuracy of 293/300 (97.7%), with significantly improved precision (31/35, 88.6%) and F-score (89.7%), demonstrating a superior ability to minimize false positives while maintaining recall at 31/34 (91.2%). In delay detection, SAFE–AI recovers from the failures of the LLM+Ontology model, improving accuracy to 274/300 (91.3%), precision to 60/78 (76.9%), and recall to 60/68 (88.2%), leading to the highest F-score of 82.2%. Confusion matrices are shown in Figure 3. These results indicate that SAFE–AI successfully integrates structured reasoning with adaptability, ensuring robustness across different medical decision-making tasks.

**Figure 3:**
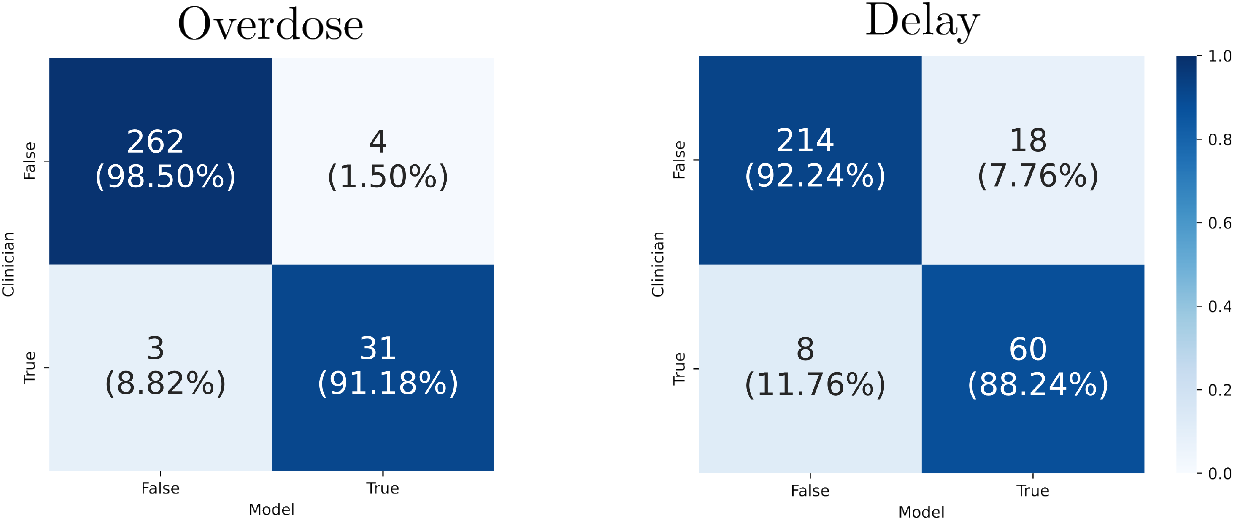
Confusion matrices for overdose and delay ASEs.

## Discussion

In this study, we demonstrated the capabilities of SAFE–AI in detecting events that are both rare and clinically important. We evaluated the performance of SAFE–AI in predicting epinephrine overdose and delay in clinical charts, and found that SAFE–AI shows high performance in predicting both, greatly surpassing baseline LLMs, especially in the case of overdoses. Baselines showed high accuracy but low precision, which supports the hypothesis that pattern-recognition models are biased toward the most likely answer but are inadequate for true detection of rare events. In contrast, SAFE–AI applied rigorous rules and performed well with both true positives and true negatives.

SAFE–AI uses LLMs and rigorous, deterministic code for the fast and reliable identification of ASEs from both free–text clinical notes and structured data sources. By providing a verified ontology and generating logical code from it, the ASE determinations made by SAFE–AI are transparent and explainable, allowing clinicians to trace the decision-making and verify every step in the process.

In spite of the advancements made by AI, manual chart reviews by clinicians remain the gold standard in ASE detection, particularly in out–of–hospital settings [5]. Focused reviews are the most accurate method, but are inadequate for timely intervention and can only be performed retrospectively. Because manual chart are time and resource intensive, only a sampling of charts are able to be reviewed, for one of the most common sources of morbidity and mortality in the US [29, 3, 30, 31, 32, 33, 34, 35, 36]. We showcase a high-performing method for fast detection of ASEs that can enable clinicians and practitioners to identify ASEs as soon as a medical record is complete, implement corrective measures and potentially reduce patient harm. SAFE-AI’s speed compared to human reviewers is crucial to scaling quality improvement and patient safety research efforts. On average, we found that specialized expert reviewers required over 10 minutes per 2-page chart review, while SAFE-AI achieved the same task in under a minute, demonstrating its potential to assist in clinical decision-making.

We note that some disagreements between the SAFE-AI predictions and the clinician reviewers were not errors made by the model. For instance, when the human reviewers encountered contradictory data, they were likely to assume that there was a documentation error in the record, whereas the model treated every data entry individually as correct or incorrect. Furthermore, human reviewers exhibited greater leniency toward rounding errors, whereas SAFE-AI applied strict precision in handling decimal points. This discrepancy led to disagreements, particularly in edge cases that fell precisely at threshold values. For instance, our threshold for severe overdose is 10 times over the recommended dose, and EMS administering exactly 10x the recommended dose due to a decimal error was a relatively common source of ASEs. A key advantage of this work is establishing a more rigorous and consistent gold standard for identifying ASEs, addressing the inherent human subjectivity that often limits current chart review methodologies [4].

As models based on pattern recognition are trained on data, they learn to output the most likely answer given the context. However, the likelihood is driven by the frequency of candidate answers within training data rather than explicit causal relationships. In medical decision–making, unlike in research or exploratory analysis, errors can have life–threatening consequences. Thus, models require a higher standard of reliability.

SAFE–AI integrates the flexibility and speed of LLMs, as well as their ability to adapt to many different unstructured formats, with strict and transparent rules and ontologies for increased reliability. This confines hallucinations and out-of-distribution inaccuracies to the information extraction step, where hallucination is rare, and enables experts to precisely review and verify model decisions.

SAFE–AI is not without limitations. The SAFE–AI framework requires the creation of an ontology and set of rules for the intermediate and output steps to be causal and not simply based on patterns in data. Condensing the combined knowledge of experts into a suitable ontology is time-intensive; future work includes generating such ontologies from databases and using expert clinicians to verify them.

While epinephrine ASEs could be detectable by other methods that don’t involve AI, AI tools incorporate data from narrative text in addition to discrete fields. Although this proof-of-concept study was limited to a low-frequency event (epinephrine administration in pediatric OHCA), this high value intervention [37] is one of the most common medical errors in the out-of-hospital setting [38] and its detection continues to rely on manual chart review and human calculation [39]. While much of the effort in machine learning research aims to tackle ill-defined problems, clinical practice can greatly benefit from the transparency and accountability of rule–following methods. Using AI to generate rules from established guidelines balances automation and safety, and is a better use of clinician time than performing reviews directly. Future work includes developing ontologies and testing the model in other areas of patient care.

## Data Availability

We are not at liberty to release the data as it is owned by a healthcare organization. However, it might be possible to be made available upon request following approvals.

## Declarations

### Funding

This work was supported by the National Heart, Lung, and Blood Institute grant number R01HL161385.

### Role of Founder

The national Heart, Lung, and Blood Institute had no role in the design and conduct of the study; collection, management, analysis and interpretation of the data; preparation, review, or approval of the manuscript; and decision to submit the manuscript for publication.

### Conflicts of Interest

The authors declare no conflicts of interest.

### Access to Data

The data used to train and evaluate the tool will not be made public as it is owned by healthcare organizations. However, it can be made available upon request following approvals.

### Ethics approval

This project was reviewed by the Committee of Clinical Investigations (CCI), the internal review board for Beth Israel Deaconess Medical Center, and approved under protocol #2022P000948. Latest amendment approved in November 5th, 2024.

## Footnotes

1 Created in BioRender. Hsieh, T. (2025) https://BioRender.com/r57y548

## Notes

### Competing Interest Statement

The authors have declared no competing interest.

### Funding Statement

Yes

### Author Declarations

The study was approved by Beth Israel Deaconess Medical Center’s Institutional Review Board as exempt

